# Transparency and Reproducibility in the Adolescent Brain Cognitive Development (ABCD) Study

**DOI:** 10.1101/2024.05.30.24308222

**Authors:** Daniel A. Lopez, Carlos Cardenas-Iniguez, Punitha Subramaniam, Shana Adise, Katherine L. Bottenhorn, Paola Badilla, Ellen Mukwekwerere, Laila Tally, Omoengheme Ahanmisi, Isabelle L. Bedichek, Serena D. Matera, Gabriela Mercedes Perez-Tamayo, Nicholas Sissons, Owen Winters, Anya Harkness, Elizabeth Nakiyingi, Jennell Encizo, Zhuoran Xiang, Isabelle G. Wilson, Allison N. Smith, Anthony R. Hill, Amanda K. Adames, Elizabeth Robertson, Joseph R. Boughter, Arturo Lopez-Flores, Emma R. Skoler, Lyndsey Dorholt, Bonnie J. Nagel, Rebekah S. Huber

## Abstract

**Background:** Transparency can build trust in the scientific process, but scientific findings can be undermined by poor and obscure data use and reporting practices. The purpose of this work is to report how data from the Adolescent Brain Cognitive Development (ABCD) Study has been used to date, and to provide practical recommendations on how to improve the transparency and reproducibility of findings.

**Methods:** Articles published from 2017 to 2023 that used ABCD Study data were reviewed using more than 30 data extraction items to gather information on data use practices. Total frequencies were reported for each extraction item, along with computation of a Level of Completeness (LOC) score that represented overall endorsement of extraction items. Univariate linear regression models were used to examine the correlation between LOC scores and individual extraction items. Post hoc analysis included examination of whether LOC scores were correlated with the logged 2-year journal impact factor.

**Results:** There were 549 full-length articles included in the main analysis. Analytic scripts were shared in 30% of full-length articles. The number of participants excluded due to missing data was reported in 60% of articles, and information on missing data for individual variables (e.g., household income) was provided in 38% of articles. A table describing the analytic sample was included in 83% of articles. A race and/or ethnicity variable was included in 78% of reviewed articles, while its inclusion was justified in only 41% of these articles. LOC scores were highly correlated with extraction items related to examination of missing data. A bottom 10% of LOC score was significantly correlated with a lower logged journal impact factor when compared to the top 10% of LOC scores (β=-0.77, 95% -1.02, -0.51; *p*-value < 0.0001).

**Conclusion:** These findings highlight opportunities for improvement in future papers using ABCD Study data to readily adapt analytic practices for better transparency and reproducibility efforts. A list of recommendations is provided to facilitate adherence in future research.

## INTRODUCTION

Transparency in science is necessary for reproducibility. Transparency pertains to the openness and accessibility of methods and analytic decisions involved in the research process (1). Despite the benefits of transparency, there exists substantial variance in the reporting of methodology. Consequently, this impacts reproducibility in science. Reproducibility refers to the minimum requirements necessary to generate the same analytic results when using a common dataset (2). Although separate concepts, many concerns related to reproducibility stem from past failed efforts related to replicability, the quality of obtaining consistent results across studies with the same research question using new data (3, 4). A landmark study examining 100 psychology papers reported that only 36% of studies reporting statistically significant findings could be replicated, and that effect sizes were routinely overestimated (5). This problem is not specific to any discipline. A reproducibility project in the cancer biology field reported a 46% replication success rate, and reproducibility hurdles included a lack of publicly available analytic code and failures to report statistical analyses in 81% and 40% of reviewed papers, respectively (6, 7). There are multiple reasons for the transparency and reproducibility problems. A survey of 1,576 researchers found that low reproducibility may be due to the unavailability of methods and code, while more statistical rigor and better mentorship emerged as practices that could improve reproducibility (8). Many of these issues are rooted in an academic culture that relies on “statistically significant” results rather than focusing on the practical relevance of findings (9, 10). The reliance on statistical thresholds can lead to questionable research practices (e.g., *p-*hacking) (11). These shortcomings in the research culture can hinder transparency efforts and compromise the public’s trust in science.

Making data available to the broader research community through an open science model promotes collaboration, replication of findings, and acceleration of scientific discovery (2). Large publicly available datasets, like the Adolescent Brain Cognitive Development (ABCD) Study^®^, have made it possible for researchers to access and analyze research data from various domains such as behavioral, genetics, and neuroimaging (12). The ABCD Study is the largest longitudinal study of brain development in children in the United States with 11,875 youth enrolled at the ages of 9 to 10 years from 21 sites that will be followed for approximately 10 years (13). This study is a critical resource for the field of developmental neuroscience that offers researchers a wealth of data to better understand factors that influence adolescent brain development. Data from the study has already been used to help chart neurodevelopmental trajectories (14) and its findings will continue to impact our current understanding of adolescent development. Furthermore, findings from the study may inform education, policy, and transform clinical practice by identifying early markers of psychopathology and substance use that can lead to targeted interventions and prevention strategies. Recent research, largely in psychology and neuroscience, has stressed the importance of transparency in methodology to improve the reproducibility of findings (15, 16). Among these recommendations include openness in sharing analytic decisions (e.g., number of excluded participants) (15–17). Whether guidance from recent initiatives has influenced the reporting of ABCD Study findings is not known.

The ABCD Study’s influential role in developmental neuroscience and adolescent health necessitates a thorough accounting of how the data have been used. The open science model embraced by the ABCD Study also provides an opportunity to assess the reporting practices in the field, identify gaps, and provide recommendations. Assessing and improving reporting practices is essential for ensuring the reliability, reproducibility, and transparency of research findings. The ABCD Study, given its extensive scope and rigorous methodologies, can play a pivotal role in elevating reporting standards in developmental neuroscience.

The aims of the current study are two-fold. The first aim was to assess the reporting practices of publications, focusing on those using the ABCD Study publicly available data due to its use by many researchers. To do so, we reviewed and extracted information related to data use practices from papers that used the ABCD Study data. The second aim was to provide a set of recommendations on best practices when using the ABCD Study data that may also be applied more broadly to the field of developmental neuroscience. The motivation to examine the data use came from the goals of the ABCD Justice, Equity, Diversity, and Inclusion (JEDI) Responsible Use of Data Workgroup. The JEDI Workgroup strives to ensure that ABCD Study data are used in a way that prevents further stigmatization or marginalization of individuals. To this end, the JEDI Workgroup focuses on creating resources for ABCD data users to encourage best practices and facilitate responsible data use.

## METHODS

A list of articles mentioning the ABCD Study were provided to the authors by the ABCD Data Analysis, Informatics & Resource Center (DAIRC). The list of articles (n=676) included full research articles, abstracts, news articles, and research letters that were published from April 2017 (the date of the first ABCD Study publication) to May 2023. All articles were obtained either through open access or via institutional membership. Articles were considered for inclusion if they were in English and peer reviewed. Abstracts and research letters were analyzed separately. News articles, reviews, commentaries, and methods papers (i.e., manuscripts that predominantly described the protocol and instruments used in the ABCD Study) were excluded.

Articles were reviewed by a team of 28 researchers (22 Research Assistants at 15 separate ABCD Study sites and 6 PhD level researchers) across the ABCD Consortium. All reviewers received training on data extraction, and the full list of measures was accompanied by a written and video tutorial that included a more in-depth explanation with visual examples (i.e., an extract from an ABCD Study paper). Each reviewer was assigned up to 25 articles. Google Forms was used for the collation of items extracted from each article reviewed, and then the submission was assessed for eligibility for inclusion in the main analysis by the first author (DAL), who then checked each Google Forms submission to determine the year of publication and whether the article was an abstract, research letter, methods, or a full-length paper. The original list of articles included some duplicates due to resubmissions (e.g., an article was resubmitted due to an erratum or retraction). This resulted in certain manuscripts being reviewed more than once. Discrepancies between duplicate submissions (e.g., due to a different response for the same question) were compared after a separate review of the manuscript by DAL. The duplicate submission with the worst overall accuracy as determined by the lead reviewer was removed from the main analysis. The overall accuracy rate between duplicate submissions was 83.9%. In total, there were 83 method papers and 15 duplicate articles excluded from the main analysis. In addition, reviewers were asked to flag any problematic articles that required further discussion. A note section was included at the end of each form where reviewers could provide any additional information that they felt was important. Flagged articles (n=8 full-length articles) and note sections were evaluated by DAL and addressed when necessary. Reviewers communicated with DAL to resolve any questions and to fix data entry errors prior to the analysis. The main analysis included only full-length manuscripts that analyzed ABCD Study data.

### Data extraction items

There were 32 items extracted from each research article, and 10 gated items that were only asked if the previous question was endorsed. The data extraction items were created after a review of the neuroimaging best practices literature (15, 16, 18). The items covered a wide range of issues related to transparency and reproducibility. Items related to replicability were not included due to the focus on the ABCD dataset. Data extraction items were initially created by the lead reviewer and were then reviewed by 5 PhD level researchers. Following feedback from the PhD level researchers, revisions were made and a final list of the data extraction items were created. The full set of data extraction items was then piloted by three RAs and two PhD level researchers. Individual items had a “Yes” or “No” option to note the presence or absence of the practice, and four measures included a third option. The full list of items can be found at: https://osf.io/qkefw/ and in the supplementary materials.

Overall, items 1 through 12 were concerned with analysis-level reproducibility, and items 13-26 broadly covered the transparent reporting of methods and results. Items 1-6 addressed the sharing of analysis scripts, software used, ABCD data release and Digital Object Identifier (DOI) information. Items 7-12 dealt with missing data issues, and whether articles considered limitations related to nonrandom missingness. Items 13-17 inquired about the inclusion of sociodemographic variables and had a gated question asking whether the article explained the inclusion of the variable as has been recommended for sound statistical modeling (19). Sociodemographic variables were emphasized due to their widespread habitual inclusion in models. Item 18 and 18b asked whether the article mentioned the manipulation of any variable (e.g., categorizing a continuous variable), and whether there was reasoning for the manipulation. Items 19-26 were concerned with the reporting of effect estimates and *p*-values, multiple comparisons, testing of statistical assumptions, and exploration of variables. Item 24 asked whether the researchers used the Data Analysis and Exploration Portal (DEAP) for their analysis. DEAP is a statistical analysis platform hosted by the DAIRC that can be used to analyze ABCD data (20). Items 27-29 were used to further identify characteristics of the article (e.g., whether imaging or genetics data were used). Items 30-32 were related to preregistration, the discussion of limitations, and whether author contributions were included.

### Level of Completeness

A Level of Completeness score was calculated for each article to summarize overall adoption of practices. Completeness has previously been used to assess adherence to recommended practices in articles analyzing large cohort studies (21). The Level of Completeness score was calculated by first converting binary variables into a numeric variable (Yes=1, No=0) and then summing up the following items: 1, 1b, 2, 2b, 3, 4, 5, 6, 7, 8, 9, 10, 11, 13b, 14b, 15b, 16b, 18b, 19, 19b, 20, 21b, 22, 23, 25, 26, 30, 31. The included items can be found in the supplementary materials (Figure S1). Items 10, 11, and 31 were recoded from a categorical to a binary variable to improve overall interpretation. Items 10 and 11 had the “no participants were excluded” responses recoded as a 0. Item 31 had a “no missing data/attrition” response that was coded as a 0 (n = 71). Item 28b was related to rationalization of brain regions when using imaging data and was not included to avoid score inflation. Item 32 (whether author contributions were indicated in the manuscript) was not included in the Level of Completeness score due to the journal specific nature of the item. The possible range of Level of Completeness scores was 0 to 28, with a higher score indicating a greater level of adoption of data extraction items.

The decision to use all items for a summary score was made after a review of the underlying structure of the data extraction items. An exploratory factor analysis (EFA) with an oblimin rotation was conducted on the data extraction items using Mplus version 8.1 (22). Binary items were treated as dichotomous variables by using the tetrachoric correlation structure. The results of the EFA (Table S1-S4) did not find evidence of a meaningful clustering between items.

### Statistical analysis

The percentage of endorsement of each item was reported for articles included in the main analysis. Percentages were calculated and reported separately for abstracts and research letters. Additional analyses used univariate linear regression to examine the correlation between individual extraction items and Level of Completeness scores. The purpose of the regression was to examine the overall influence of each extraction item on Completeness score. Results of the univariate linear regression represent the score difference associated with individual items. Statistical analysis of the data was performed using R version 4.2.2 and R Studio version 2023.06.01 build 524 (23, 24).

Figures were created using the ggplot2 package in R (25). Checks for normality were performed using the car package in R (26). Data preparation was completed using Microsoft Excel and the dplyr package in R (27, 28). R scripts are available at: <u>github.com/Daniel-Adan-Lopez/ABCD_Transparency_Reproducibility</u>.

### Sensitivity analysis

A machine learning wrapper algorithm was used to estimate variable importance in relation to Level of Completeness score via feature selection. With the Boruta package (version 8) in R, we used a Random Forest based feature selection method to determine important and non-important attributes (29). A Boruta model was created using Level of Completeness score as the dependent variable and the data extraction items as the independent variable (i.e., features). Missing values were coded as “Not applicable” to prevent errors while running the Boruta wrapper algorithm.

### Post hoc analysis

A post hoc analysis was conducted to examine the relationship between Level of Completeness score and journal impact factor. The most recently disseminated 2-year journal impact factor was collected for articles that scored in the top 10% (n=56), middle 10% (n=56), and bottom 10% (n=56) of Completeness scores. Only full-length articles (i.e., not abstracts, methods papers, or research letters) were included in the post hoc analysis. The impact factor was logged to reduce the influence of outliers (Figure S2). A univariate linear regression model was then used to examine the correlation between Completeness category (bottom 10%, middle 10%, top 10%) and the log of the impact factor. Completeness score was also examined as a continuous measure.

## RESULTS

The initial review included 679 articles. Abstracts (n=18) and research letters (n=11) were separately analyzed. An additional two articles were removed due to not analyzing ABCD data. The final analysis included 549 full-length research articles (Figure 1). Percentages of each response for the data extraction items can be found in Figure 2.

**Figure 1.**
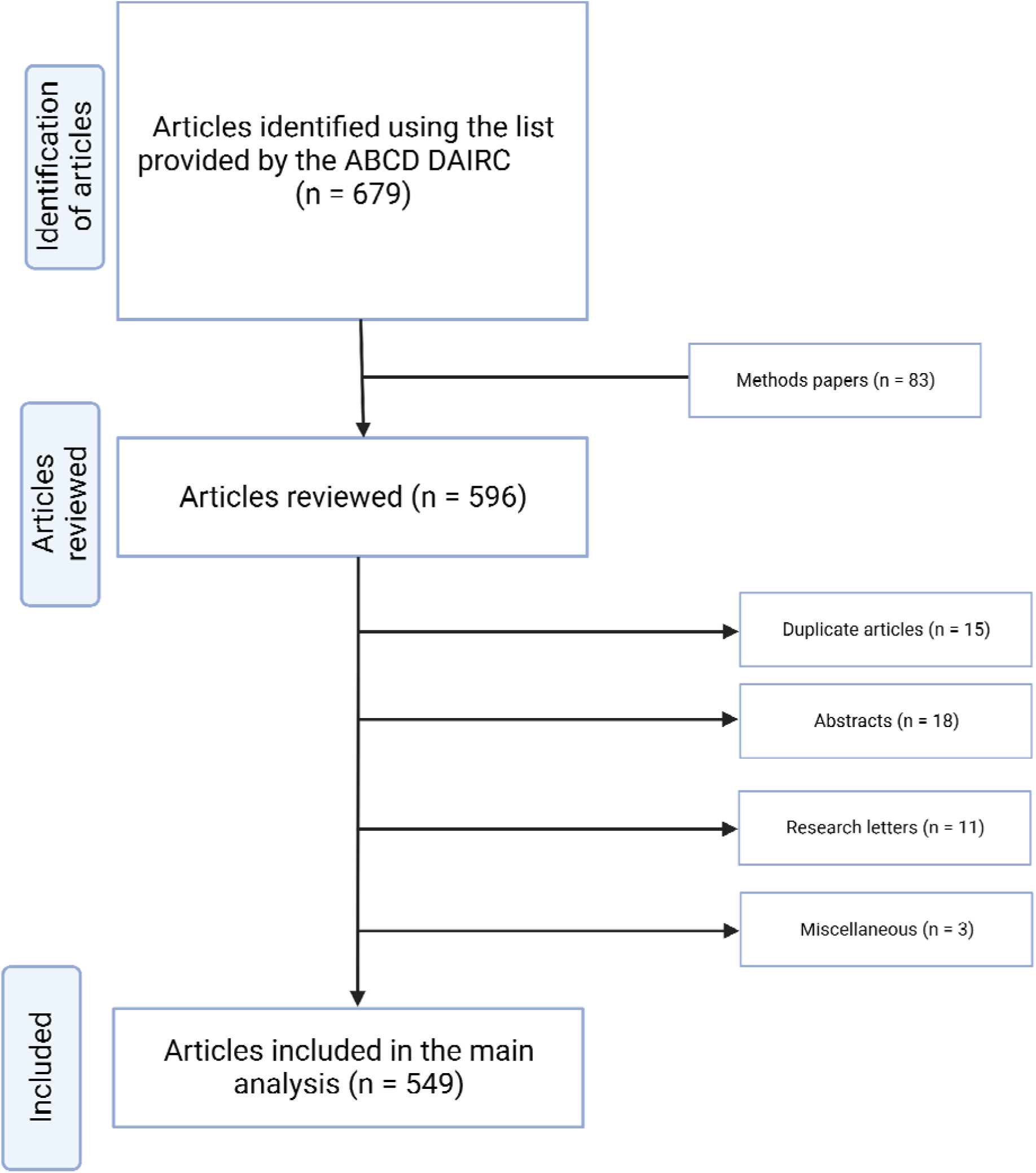
Flow chart summary of the article screening process

**Figure 2.**
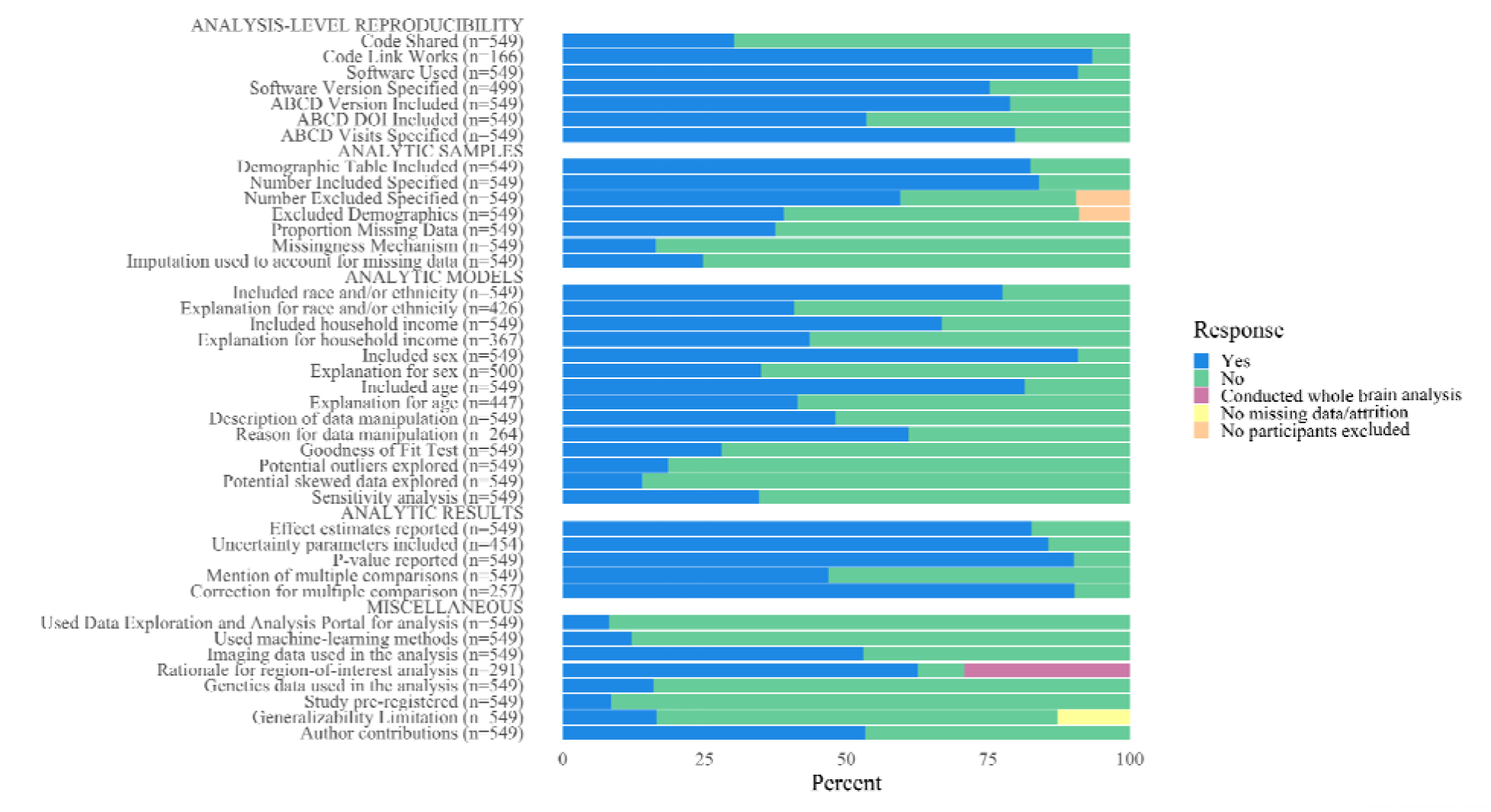
The percentage of response types for each data extraction item

### Analysis-level reproducibility

A link to the analysis scripts were included in 30.2% (n=166) of the articles, and 93.4% (n=155) of the included links were active and working. Software information was mentioned in 91% (n=499) of the articles. The software version (e.g., R version 4.2.2) was included in 75.3% (n=376) of articles. The ABCD Study data release version (e.g., ABCD version 4.0) and digital-object identifier (DOI) were mentioned in 79% (n=433) and 54% (n=294) of articles, respectively. The ABCD Study visit number (e.g., Baseline, 1-year follow-up visit) was explicitly mentioned in 79.8% (n=438) of articles.

### Analytic Samples

A table describing the study sample was included in 82.5% of articles (n=453). The sample size of the final analytic model was mentioned in 84% of articles (n=461), and 59.6% of articles (n=327) mentioned the number of participants excluded from the final analyses due to missing data and/or exclusion criteria. The characteristics of the excluded sample were detailed in 42.8% of papers that reported missingness in the data. Quantification of missing data for individual variables (e.g., the percent missing household income data) was included in 37.5% (n=206) of articles. The missing data mechanism (e.g., missing completely at random) was discussed in 16.4% (n=90) of articles. Imputation methods were used to account for missing data in 24.8% (n=136) of articles, and in 36.9% (n=76) of papers that mentioned missing data for individual variables. Imputation methods were also used in 61.1% (n=55) of papers that examined the missingness mechanisms (e.g., missing completely at random).

### Analytic Models

Race and/or ethnicity was included in 77.6% of papers (n=426), and its inclusion was explained in 40.8% of these articles (n=174). Household income was included in 66.8% of papers (n=367), and its inclusion was explained in 43.6% of these articles (n=160). Participant sex was included in 90.9% (n=499) of reviewed articles, and its inclusion was explained in 35% of these articles (n=175). Participant age was included and explained in 81.4% (n=447) and 41.4% (n=185) of articles, respectively.

Data manipulation (e.g., changing a variable from continuous to categorical) was mentioned in 48.1% (n=264) of reviewed articles, and was explained in 61% of these articles (n=161). Effect estimates (e.g., a beta coefficient or Cohen’s d) were reported in 82.7% (n=454) of articles included in the main analysis. A quantification of uncertainty (e.g., confidence intervals, standard errors) was included in 85.7% (n=389) of articles that reported an effect estimate. A *p*-value was reported in 90.2% (n=495) of articles included in the main analysis.

Multiple comparisons were mentioned in 46.8% of articles (n=257), and correction for multiple comparisons was detailed in 90% (n=232) of these articles. A sensitivity analysis was conducted in 34.6% (n=190) of articles included in the main analysis. A goodness of fit test (e.g., Akaike Information Criterion) to select the final model was mentioned in 28.1% (n=154) of articles. Potential outliers and skewed data were discussed in 18.6% (n=102) and 14% (n=77) of articles, respectively.

### Miscellaneous

Imaging data were used in 53% (n=291) of articles included in the main analysis, and 92% (n=267) of these articles included a rationale for selecting certain regions of interest. Genetics data were analyzed in 16% (n=89) of articles. Imaging and genetics data were included in 9.1% of articles (n=50). Machine-learning methods were used in 12.2% of articles included in the main analysis (n=67), and 76.1% (n=51) of these also used imaging data. The Data Exploration and Analysis Portal (DEAP) was used to analyze ABCD Study data in 8.2% (n=45) of articles included in the main analysis. A statement about limitations to generalizability due to missing data was included in 28.6% of articles that reported at least some missing data (n=69). Author contributions were included in 53.4% of articles (n=293). Study preregistration was mentioned in 8.6% (n=47) of articles included in the main analysis.

### Level of Completeness score

The Level of Completeness score ranged from 1 to 23 in full-length articles(Figure 3). The mean score was 13.11 (median = 13.0, standard deviation [SD] = 3.92, variance = 15.4). The Level of Completeness score in abstracts (n=18) ranged from 0 to 13 (mean = 3.7, SD = 2.9). The range of scores in research letters (n=11) was 0 to 12 (mean = 8.64, SD=3.2).

**Figure 3.**
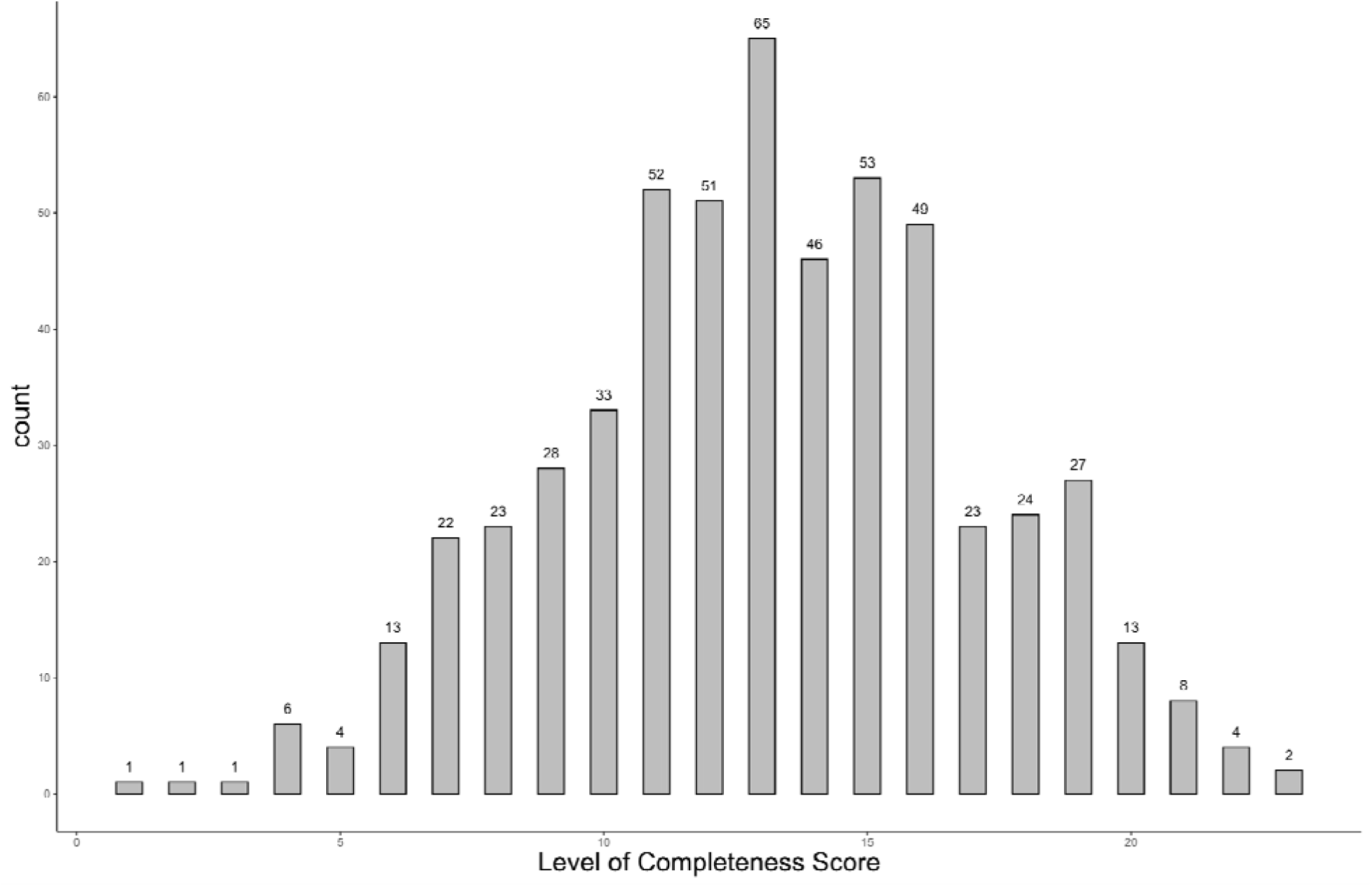
The frequency and distribution of Level of Completeness score in the articles included in the main analysis

Results of the univariate linear regression with individual data extraction items emphasize the importance of missing data (Table 1). Specifying the total number of participants excluded due to missing data was significantly correlated with Level of Completeness scores (β = 4.17, 95% Confidence Interval [CI]: 3.6, 4.8; *p*-value <0.0001). A description of missingness for individual variables (e.g., household income) was positively correlated with Completeness scores (β = 4.1, 95% CI: 3.5, 4.7; *p*-value = <0.0001). Mention of the ABCD data version and ABCD Study visits was significantly correlated with higher Completeness scores (β = 3.8 and β = 3.4, respectively). Using the DEAP was correlated with a decreased Completeness score (β = -1.9, 95% CI: -3.1, -0.7; *p*-value = 0.017). There was no significant difference in Completeness scores in studies that used machine learning methods (*p*-value = 0.38). The year of publication was positively correlated with Level of Completeness score (β=0.83, 95% CI: 0.55, 1.11; *p*-value <0.0001). In other words, more recent publications had higher Completeness scores. Results of all extraction items can be found in the supplementary materials (Table S5).

**Table 1.**
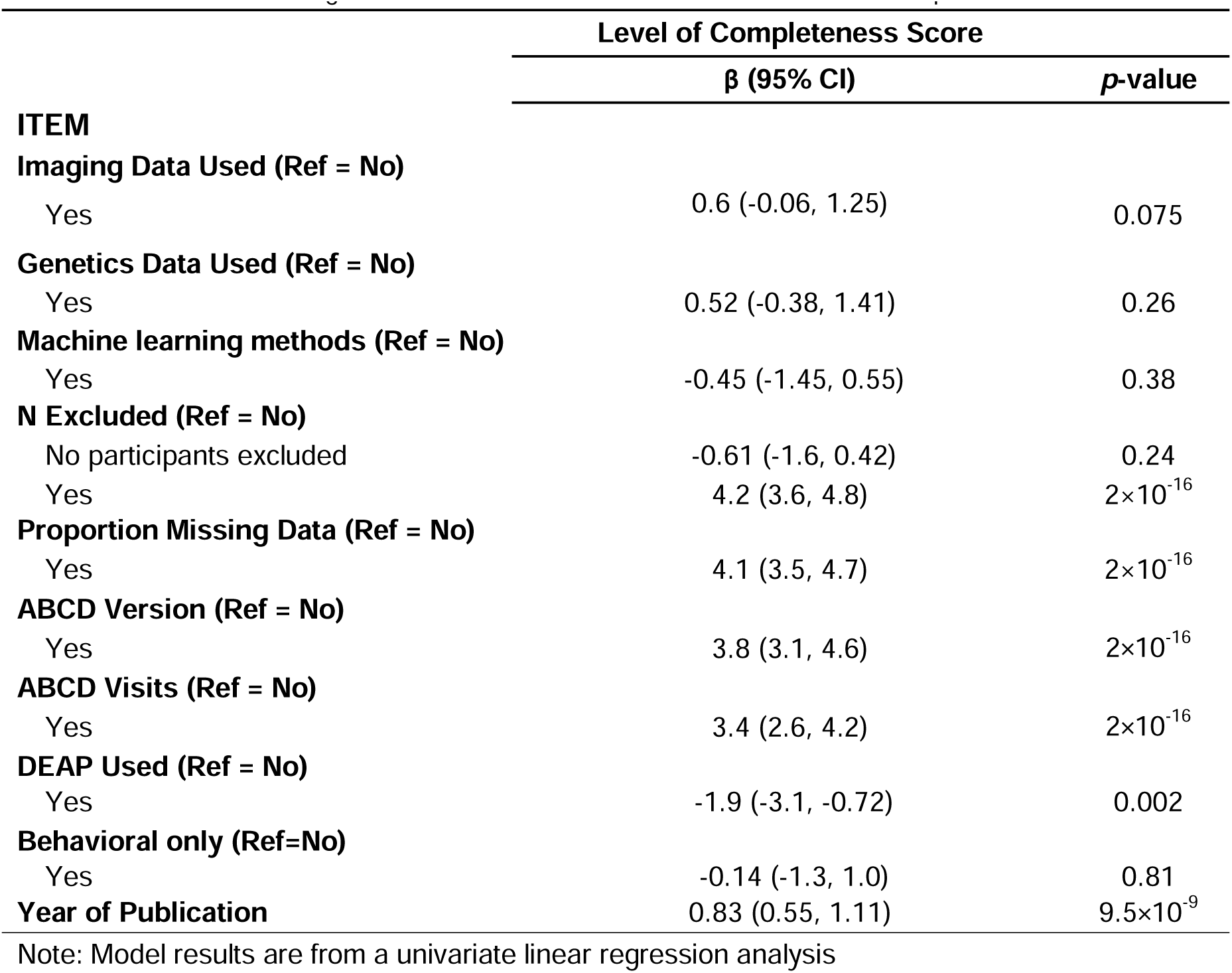
Univariate linear regression results with extraction items and Level of Completeness score.

### Results of the sensitivity analysis

Findings from the sensitivity analysis were like those found using univariate linear regression. Mentioning the number of participants excluded from the final analytic model (i.e., item #8) was the most important predictor of Level of Completeness score (Figure 4). The following items were considered nonimportant contributors of Completeness score: Machine Learning and Author Contributions. The full Boruta attribute statistics can be found in Table S6.

**Figure 4.**
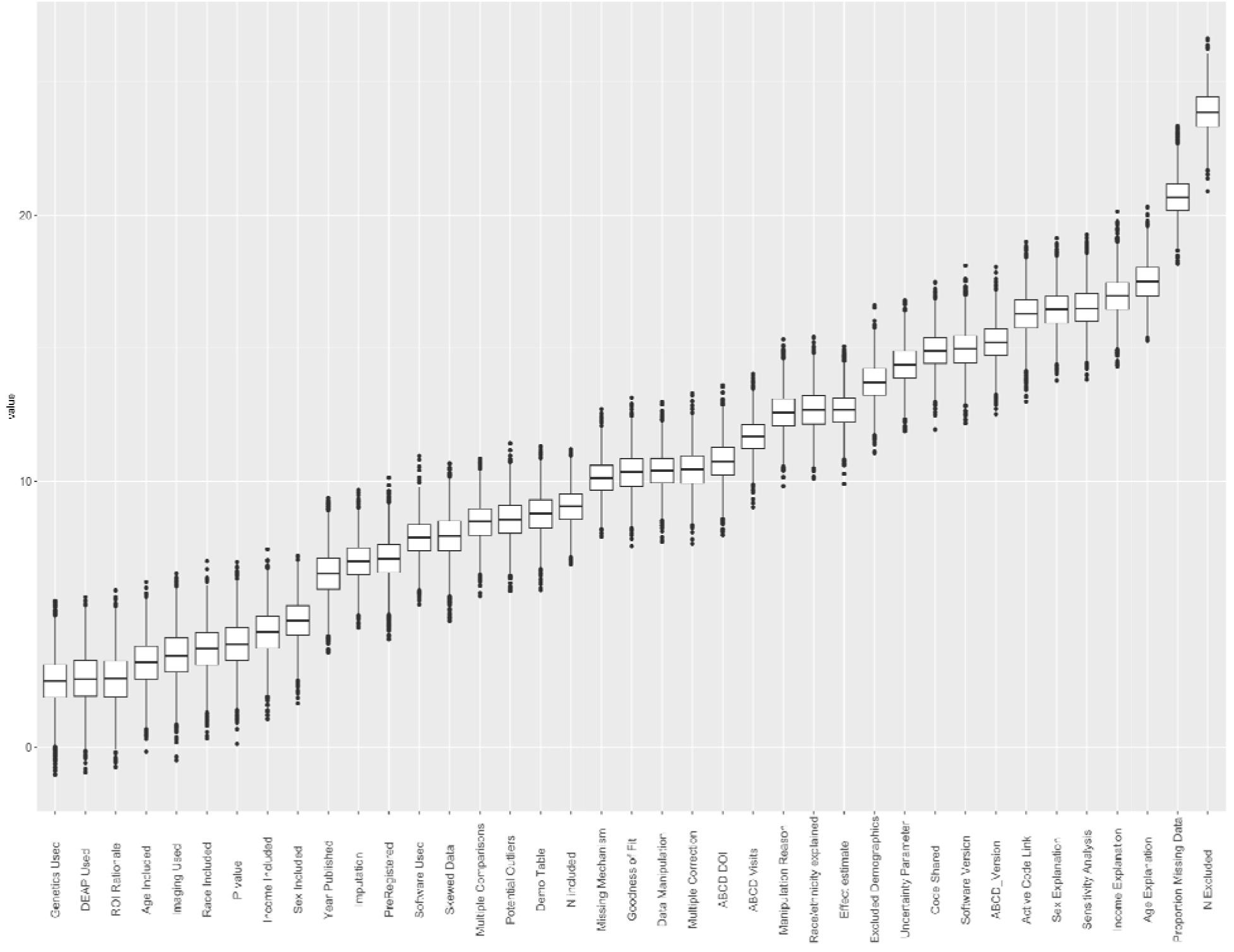
Results of the machine learning feature selection method

### Post hoc analysis

The post hoc analysis examined the relationship between Level of Completeness score and the logged journal impact factor (Supplementary Figure S3). The Pearson correlation coefficient between the Level of Completeness score and logged journal impact factor was 0.43. There was 1 article in the top 10%, 5 articles in the middle 10%, and 4 articles in the bottom 10% of Completeness scores that were in journals without an impact factor. The mean 2-year journal impact factor of the included articles (n=158) was 6.9 (range = 0.5 to 82.9). The mean impact factor for the bottom 10%, middle 10%, and top 10% of Completeness scores was 3.9, 7.2, and 9.5, respectively. A bottom 10% Completeness score was significantly correlated with a 0.77-point lower logged impact factor (*p*-value < 0.0001) when compared to articles in the top 10% of Completeness scores (Table S6). There was no significant difference in the logged impact factor of manuscripts in the top 10% and middle 10% of Completeness scores (*p*-value=0.54). Completeness score was significantly correlated with the logged impact factor in models using Completeness as a continuous measure (β=0.056, 95% CI: 0.04, 0.07; *p*-value<0.0001). In other words, a one-point increase in the Completeness score was correlated with a 0.06-point increase in the logged impact factor.

## DISCUSSION

The findings highlight shortcomings and optimistic reporting trends in the level of analytic information included in ABCD Study publications, with implications for transparency and reproducibility. Analytic scripts were shared in fewer than a third of publications included in the main analysis, and this proportion increased to 44% of papers published in 2023. Issues related to missing data were intermittently addressed. The sociodemographic differences of participants included and excluded from the analysis were provided in fewer than half of publications. Descriptions of missingness for individual variables were included in around one-third of articles overall. The reasons for inclusion of sociodemographic variables (e.g., race and/or ethnicity, household income) was explained in fewer than half of the papers. Around 20% of articles mentioned problems with generalizability of results due to nonrandom missingness (e.g., disproportionate missing data/attrition in minority groups). Univariate regression models and machine learning methods highlighted missingness items as an important contributor of Level of Completeness scores. Finally, the post hoc analysis found a significant positive correlation between Level of Completeness scores and journal impact factor. Together, these findings highlight existing gaps in the statistical rigor and reporting of studies using ABCD Study data but indicate that some of these gaps are already being closed by improved reporting practices in recent years. Based on the study findings, we have provided recommendations (summarized in Table 2) for best reporting practices to improve transparency and reproducibility for future full-length ABCD Study articles. The recommendations are tailored to the ABCD Study but can be applied to other observational data in the field of developmental neuroscience.

**Table 2.** List of practices and recommendations to improve transparency and reproducibility in ABCD Study publications.

| **Practice** | **Recommendations** |
| --- | --- |
| **1. Code sharing** | Make analytic scripts publicly available on an open platform (e.g., GitHub, OSF). The scripts should include enough information to reproduce the findings of the study. Add detailed comments to each step of the code to improve interpretability. |
| **2. Software Used** | Clearly state the software and software version that was used for all steps of the analysis. |
| **3. ABCD Version** | State which ABCD data version is being used in the analysis (e.g., ABCD version 4.0). |
| **4. ABCD Visits** | State the ABCD Study visits that were included in the analysis. |
| **5. Demographics Table** | Include a table in the main paper or supplementary materials detailing the characteristics of the analytic sample. |
| **6. Included Sample Size** | State the final sample size of your analytic model after any exclusions. |
| **7. Excluded Sample Size** | State the number of participants that were excluded. |
| **8. Excluded Demographics** | Include a table in the main paper or supplementary materials detailing the characteristics of the excluded sample. |
| **9. Missing data - covariates** | Describe the amount of missing data in the variables included in your analysis. |
| **10. Missing data - attrition** | Describe the amount of missing data due to a participant not attending a study visit. |
| **11. Covariate explanation** | Provide a reason for including a covariate in the model for adjustment. The reason should include support for why it was necessary (e.g., a citation of previous research or meets the definition of a confounder). |
| **12. Data Manipulation** | Describe and include a reason for variable manipulation. For example, justify why a continuous measure was categorized, and why certain cut-offs were used. |
| **13. Effect estimates** | Include a measure of association along with quantification of uncertainty (e.g., confidence intervals, standard errors). |
| **14. Sensitivity Analysis** | Test whether the results are robust to any statistical assumptions. Some common assumptions include how a variable is defined or that the results are robust to unmeasured confounding. |
| **15. Skewed Data** | Examine skewness in the variables of interest to determine whether it meets model assumptions. Explore alternatives if model assumptions are not met (e.g., generalized linear models, outcome transformation). |
| **16. Potential Outliers** | Examine any potential outlying data points in your covariates. Decide whether these were due to measurement error or natural variability in the measurement. |
| **17. Rationale for ROIs** | Provide the rationale and method for selecting the particular regions of interest when analyzing imaging data. Explicitly detail any additional steps taken when analyzing the imaging data (e.g., regressing out motion parameters, adjustment for scanner). |
| **18. Study preregistration** | Preregister the study hypothesis and analytic plans prior to analyzing the ABCD data. Preregistration can help guard against publication bias and P-hacking. Preregistration can be done on open platforms like the Open Science Framework (OSF). Include a link of the preregistration in your manuscript. |
| **19. Limitations** | Describe any caveats to the findings. For example, that the generalizability of the findings may be impacted by nonrandom attrition. |

### Analysis-level Reproducibility

Analytic scripts that detail how the data were processed and manipulated can create support for scientific claims (30). Code sharing reluctance may be due in part to a lack of awareness. A survey by Stuart et al. of over 7,700 researchers found that 33% did not know where to deposit data and code, and 46% were unsure how to usefully organize the data (31). Reproducibility issues are heightened by the lack of code sharing. A case study comparing reproducibility before and after a journal policy change reported a 40% increase in the probability of reproducing results when analytic code was made available (32). In that study, only 37% of papers that provided data, but not code, were reproducible compared to 81% of papers that included analytic scripts (32). Altogether, this highlights the need for greater adoption of code sharing practices in the field of developmental neuroscience to improve transparency and reproducibility efforts.

However, the current study found that publications from 2023 (43.3% shared code compared to 29% in 2022 and 30% in 2021) were more likely to share analytic scripts and highlighted the recent emphasis on improving transparency and reproducibility efforts. The ABCD Study affords an incredible opportunity to improve current code sharing practices as it provides resources for researchers interested in analyzing the ABCD dataset. For example, the ABCD-ReproNim (http://www.abcd-repronim.org) course was designed to improve reproducibility efforts and is still available and free to use at a self-guided pace. Additionally, publicly available analytic scripts on platforms like GitHub can be used by any researcher to determine which variables, methods, and assumptions were made while working with the open source ABCD data. Code sharing can also be used to reanalyze the data with alternative methods that can lead to new insights. The burden of providing well-documented code may deter some researchers from sharing analytic scripts, although even poorly documented code can be beneficial to improving reproducibility efforts in science (33, 34). Future ABCD data use agreements should consider requiring code sharing on public platforms (e.g., GitHub, Open Science Framework) to normalize transparency efforts. Based on our findings, we strongly recommend widespread sharing of analytic scripts that can be used to reproduce ABCD Study findings. The analytic scripts should, at minimum, include publicly accessible code that can recreate research results (e.g., statistical models). We highly encourage the inclusion of comments throughout the analytic script to improve interpretability. In addition to sharing code, we encourage researchers to share version of ABCD data and the visits included in the analyses as well as the software and version used for all steps of the analysis.

### Analytic Samples

Most papers using ABCD data included a description of the study sample. The high percentage is in line with previous findings of participant characteristics reporting in neuroimaging studies (35). In contrast, issues related to missing data were not regularly addressed in ABCD papers. Missing data can introduce bias and weaken the generalizability of findings when the missingness is nonrandom (36, 37). These data and previous studies exploring missingness in research suggests that the rate of missingness reporting in papers using ABCD Study data is in line with that of the broader literature. A study of psychology papers (n=113) published in 2012 found that 56% of papers mentioned missing data, and 6.7% explored how missingness was related to other variables (38). A review of cohort studies (n=82) found that 43% reported the amount of missing data at follow-up visits, and that 32% compared the characteristics of participants and nonparticipants (39). A separate review of developmental psychology papers (n=100) found that only 43% of studies that mentioned missing data compared the two groups (40).

Missing data in the ABCD Study can arise from selected exclusion criteria (e.g., failed imaging quality control, refusal to answer) or from attrition (e.g., loss to follow-up) (41). In the case of ABCD imaging data, exclusion criteria can result in nonrandom missingness that is highly correlated with sociodemographic variables. For example, more than 50% of Black children that completed the EN-back and Stop Signal fMRI task during the baseline ABCD visit were excluded after data processing (42). The current study also found that techniques for dealing with missing data (e.g., multiple imputation) were used infrequently. The decision to use imputation methods depends on many different factors related to the study (e.g., the amount of missing data, the missingness mechanism). Imputation may be necessary when there is nonrandom or systematic missingness of data (43). Caution is warranted when considering imputation since it does not always reduce bias (44). Altogether, there is considerable evidence that missing data is not being sufficiently addressed in papers using ABCD Study data. Future studies using the ABCD data should examine patterns of missingness and make a greater effort to describe the population excluded from the analytic sample. We recommend that data users state the final sample size after exclusions, include the number of participants excluded, and include a table that details the characteristics of the excluded sample. Researchers should also examine how missing data is correlated with their outcome variable and/or other covariates, since this may be an indicator of whether the missingness is ignorable (45). For example, a researcher conducting a longitudinal analysis using the Child Behavior Checklist – Externalizing subscale as their outcome of interest can describe participants with and without a missing ABCD time point. A significant difference in the characteristics of the two groups can be an indicator of selection bias and should be mentioned as a potential limit to generalizability.

### Analytic Models

The current study found that sociodemographic variables were commonly included in statistical models without explanation. This finding is in line with previous reports from other studies. A study using randomly selected articles (n=60) from psychology journals found that 18.3% of articles failed to provide an explanation for any control variables, and 53.3% for at least one variable (46). A study of Management Research journals found that 19% of publications (n=162) did not provide full justification for variable control (47). Another study of Management Research publications (n=812) from 2005 to 2009 found that 18.2% of articles provided no rationale for variable inclusion (48).

The current study looked for any explanation for variable inclusion (e.g., a confounder, a citation supporting the inclusion). There are statistical and social reasons for justifying the inclusion of sociodemographic variables. Controlling for certain variables (e.g., a mediator) can inadvertently result in biased effect estimates and may even induce spurious associations (e.g., adjustment for a collider) (49, 50). More thoughtful consideration of variable relationships (e.g., using directed acyclic graphs) can help determine whether statistical adjustment is necessary (51). The habitual inclusion of sociodemographic variables (e.g., race and/or ethnicity) in statistical models has also been criticized for perpetuating stigmatization and inequity (52). We strongly encourage that researchers provide support for the inclusion of all covariates in their statistical models. This practice can alleviate concerns over biased estimates due to improper statistical adjustment (e.g., conditioning on a collider). In particular, the use of any race and/or ethnicity variable should be thoroughly explained and justified. Researchers should determine whether inclusion, omission, or some other method (e.g., effect measure modification) is necessary when considering the use of race or ethno-racial variables. Researchers should consult recommended best practices (51–53) prior to using race and/or ethnicity variables in their analyses.

Finally, the ABCD dataset includes hundreds of socioeconomic and environmental variables that may offer superior explanatory power than sociodemographic variables. We encourage exploration of the ABCD data dictionary (https://data-dict.abcdstudy.org/) to determine whether there are variables that are more direct measures of differences in the study sample.

### Strengths

The study had several strengths. First, we provided a comprehensive summary of reporting practices across papers using ABCD Study data. Since the ABCD Study is ongoing, the findings will hopefully influence the inclusion of important methodological and analytic information in future publications using the data, increasing transparency and reproducibility. Second, we have provided the full list of items as a resource for researchers to incorporate into their own manuscripts. A list of practices and recommendations is available (Table 2 or at OSF: https://osf.io/8wqft) and will ideally improve adherence to recommended best practices when using ABCD data and can also be applied more broadly to the field of developmental neuroscience. Third, we examined variable importance and quantified the importance of certain practices (e.g., exploration of missing data) to overall methodological and reporting rigor. We are hopeful that future work will investigate patterns of missingness in the ABCD Study dataset.

### Limitations

There were several limitations in the study. First, the extraction items were created for use with the ABCD Study data and may not generalize outside the study, although we expect that certain items and the questions motivating their inclusion will have broad applicability. Second, the extraction items were heavily influenced by neuroimaging best practices recommendations and may not have captured important measures for other fields (e.g., genetics). Third, the large number of reviewers and papers makes it difficult to feasibly address any potential measurement error or subjectivity in reviews, and the potential impact thereof on findings reported here. We cannot rule out that errors were made in the extraction of data from each article. We expect that the large sample of papers that were included in the analysis minimized the influence that measurement error had on our findings. Fourth, the absence of underlying groupings in the extraction items meant that items were summed and treated equally for the Completeness score. We expect that certain items (e.g., code sharing) are more valuable contributors to transparency and reproducibility efforts.

## Conclusion

The findings highlight glaring transparency issues in studies using ABCD data. The ABCD Study will influence many aspects of research involving adolescent development and ensuring that findings are reproducible is highly feasible and crucial to moving the field forward. We hope that our review of ABCD use of data sheds light on the many opportunities for improvement in research practices and strengthens reproducibility efforts going forward.

## Data Availability

All data produced are available online at https://osf.io/qkefw/ and github.com/Daniel-Adan-Lopez/ABCD_Transparency_Reproducibility

https://github.com/Daniel-Adan-Lopez/ABCD_Transparency_Reproducibility

## Author Contributions

**Conceptualization:** Daniel A. Lopez and Rebekah S. Huber. **Formal analysis:** Daniel A. Lopez. **Investigation:** Daniel A. Lopez, Rebekah S. Huber, Carlos Cardenas-Iniguez, Punitha Subramaniam, Shana Adise, Katherine L. Bottenhorn, Paola Badilla, Ellen Mukwekwerere, Laila Tally, Omoengheme Ahanmisi, Isabelle L. Bedichek, Serena D. Matera, Gabriela M. Perez-Tamayo, Nicholas Sissons, Owen Winters, Anya Harkness, Elizabeth Nakiyingi, Jennell Encizo, Zhuoran Xiang, Isabelle G. Wilson, Allison N. Smith, Anthony R. Hill, Amanda K. Adames, Elizabeth Robertson, Joseph R. Boughter, Arturo Lopez-Flores, Emma R. Skoler, and Lyndsey Dorholt. **Writing - original draft:** Daniel A. Lopez. **Writing - review & editing:** Daniel A. Lopez, Rebekah S. Huber, Carlos Cardenas-Iniguez, Punitha Subramaniam, Shana Adise, Anya Harkness, Anthony R. Hill, and Bonnie J. Nagel.

## Acknowledgements

We would like to acknowledge the contributions of the ABCD JEDI Responsible Use of Data Workgroup in providing feedback throughout the process of creating this paper and the ABCD JEDI Advisory Council for their support. We would like to thank the following individuals for helping with the recruitment of reviewers: Calen Smith and Laura Zeimer. We would also like to thank Rachel Weistrop at the DAIRC for providing the list of ABCD publications.

## References

1. Korbmacher M, Azevedo F, Pennington CR, et al. The replication crisis has led to positive structural, procedural, and community changes. Communications Psychology 2023;1(1):3.

2. Parsons S, Azevedo F, Elsherif MM, et al. A community-sourced glossary of open scholarship terms. Nature Human Behaviour 2022;6(3):312–8.

3. Ioannidis JP. Why most published research findings are false. PLoS medicine 2005;2(8):e124.

4. Sciences NAo, Policy, Affairs G, et al. Reproducibility and replicability in science. National Academies Press; 2019.

5. Collaboration OS. Estimating the reproducibility of psychological science. Science 2015;349(6251).

6. Errington TM, Mathur M, Soderberg CK, et al. Investigating the replicability of preclinical cancer biology. Elife 2021;10:e71601.

7. Errington TM, Denis A, Perfito N, et al. Challenges for assessing replicability in preclinical cancer biology. eLife 2021;10:e67995.

8. Baker M. 1,500 scientists lift the lid on reproducibility. Nature 2016;533(7604):452-4.

9. Joober R, Schmitz N, Annable L, et al. Publication bias: what are the challenges and can they be overcome? J Psychiatry Neurosci 2012;37(3):149–52.

10. Nuzzo R. Statistical errors. Nature 2014;506(7487):150.

11. Andrade C. HARKing, cherry-picking, p-hacking, fishing expeditions, and data dredging and mining as questionable research practices. The Journal of clinical psychiatry 2021;82(1):25941.

12. Volkow ND, Koob GF, Croyle RT, et al. The conception of the ABCD study: From substance use to a broad NIH collaboration. Developmental Cognitive Neuroscience 2018;32:4–7.

13. Garavan H, Bartsch H, Conway K, et al. Recruiting the ABCD sample: Design considerations and procedures. Developmental cognitive neuroscience 2018;32:16–22.

14. Bethlehem RAI, Seidlitz J, White SR, et al. Brain charts for the human lifespan. Nature 2022;604(7906):525–33.

15. Klapwijk ET, van den Bos W, Tamnes CK, et al. Opportunities for increased reproducibility and replicability of developmental neuroimaging. Developmental Cognitive Neuroscience 2021;47:100902.

16. Nichols TE, Das S, Eickhoff SB, et al. Best practices in data analysis and sharing in neuroimaging using MRI. Nature neuroscience 2017;20(3):299–303.

17. Munafò MR, Nosek BA, Bishop DV, et al. A manifesto for reproducible science. Nature human behaviour 2017;1(1):1–9.

18. Gilmore RO, Diaz MT, Wyble BA, et al. Progress toward openness, transparency, and reproducibility in cognitive neuroscience. Annals of the New York Academy of Sciences 2017;1396(1):5–18.

19. Greenland S, Pearce N. Statistical foundations for model-based adjustments. Annual review of public health 2015;36:89–108.

20. Heeringa SG, Berglund PA. A guide for population-based analysis of the Adolescent Brain Cognitive Development (ABCD) Study baseline data. BioRxiv 2020.

21. Gibson MJ, Spiga F, Campbell A, et al. Reporting and methodological quality of studies that use Mendelian randomisation in UK Biobank: a meta-epidemiological study. BMJ Evidence-Based Medicine 2023;28(2):103–10.

22. Muthén B, Muthén L. Mplus. Handbook of item response theory: Chapman and Hall/CRC, 2017:507–18.

23. Team R. RStudio: integrated development environment for R. 2015. 2019.

24. Team RC. R: A language and environment for statistical computing. 2013.

25. Wickham H. ggplot2. Wiley interdisciplinary reviews: computational statistics 2011;3(2):180–5.

26. Fox J, Weisberg S, Adler D, et al. Package ‘car’. Vienna: R Foundation for Statistical Computing 2012;16(332):333.

27. Wickham H, François R, Henry L, et al. Package ‘dplyr’. A Grammar of Data Manipulation R package version 2019;8.

28. Corporation M. Microsoft Excel. 2018.

29. Kursa MB, Rudnicki WR. Feature selection with the Boruta package. Journal of statistical software 2010;36:1–13.

30. Baker M. Why scientists must share their research code. Nature 2016.

31. Stuart D, Baynes G, Hrynaszkiewicz I, et al. Practical challenges for researchers in data sharing. 2018.

32. Laurinavichyute A, Yadav H, Vasishth S. Share the code, not just the data: A case study of the reproducibility of articles published in the Journal of Memory and Language under the open data policy. Journal of Memory and Language 2022;125:104332.

33. Gorgolewski KJ, Poldrack RA. A practical guide for improving transparency and reproducibility in neuroimaging research. PLoS biology 2016;14(7):e1002506.

34. Barnes N. Publish your computer code: it is good enough. Nature 2010;467(7317):753-.

35. Sterling E, Pearl H, Liu Z, et al. Demographic reporting across a decade of neuroimaging: a systematic review. Brain Imaging Behav 2022;16(6):2785–96.

36. Dong Y, Peng CY. Principled missing data methods for researchers. Springerplus 2013;2(1):222.

37. Lash TL, VanderWeele TJ, Haneause S, et al. Modern epidemiology. Lippincott Williams & Wilkins; 2020.

38. Little TD, Jorgensen TD, Lang KM, et al. On the joys of missing data. Journal of pediatric psychology 2014;39(2):151–62.

39. Karahalios A, Baglietto L, Carlin JB, et al. A review of the reporting and handling of missing data in cohort studies with repeated assessment of exposure measures. BMC Medical Research Methodology 2012;12(1):96.

40. Jeličić H, Phelps E, Lerner RM. Use of missing data methods in longitudinal studies: The persistence of bad practices in developmental psychology. Developmental Psychology 2009;45(4):1195–9.

41. Ewing SWF, Dash GF, Thompson WK, et al. Measuring retention within the adolescent brain cognitive development (ABCD) SM study. Developmental Cognitive Neuroscience 2022;54:101081.

42. Chaarani B, Hahn S, Allgaier N, et al. Baseline brain function in the preadolescents of the ABCD Study. Nature neuroscience 2021;24(8):1176–86.

43. Kleinke K, Reinecke J, Salfrán D, et al. Applied multiple imputation: Advantages, pitfalls, new developments and applications in R. Springer Nature; 2020.

44. Twisk J, de Boer M, de Vente W, et al. Multiple imputation of missing values was not necessary before performing a longitudinal mixed-model analysis. Journal of Clinical Epidemiology 2013;66(9):1022–8.

45. Twisk JW. Applied longitudinal data analysis for epidemiology: a practical guide. cambridge university press; 2013.

46. Becker TE. Potential problems in the statistical control of variables in organizational research: A qualitative analysis with recommendations. Organizational research methods 2005;8(3):274–89.

47. Carlson KD, Wu J. The Illusion of Statistical Control: Control Variable Practice in Management Research. Organizational Research Methods 2011;15(3):413–35.

48. Atinc G, Simmering MJ, Kroll MJ. Control variable use and reporting in macro and micro management research. Organizational Research Methods 2012;15(1):57–74.

49. Schisterman EF, Cole SR, Platt RW. Overadjustment bias and unnecessary adjustment in epidemiologic studies. Epidemiology (Cambridge, Mass) 2009;20(4):488.

50. Wysocki AC, Lawson KM, Rhemtulla M. Statistical Control Requires Causal Justification. Advances in Methods and Practices in Psychological Science 2022;5(2):25152459221095823.

51. VanderWeele TJ, Robinson WR. On the causal interpretation of race in regressions adjusting for confounding and mediating variables. Epidemiology 2014;25(4):473–84.

52. Cardenas-Iniguez C, Gonzalez MR. “We controlled for race and ethnicity…” Considerations for the use and communication of race and ethnicity in neuroimaging research 2023.

53. Martinez RAM, Andrabi N, Goodwin AN, et al. Conceptualization, operationalization, and utilization of race and ethnicity in major epidemiology journals, 1995–2018: A systematic review. American Journal of Epidemiology 2023;192(3):483–96.

