## Supplementary Materials for "Transparency and Reproducibility in the Adolescent Brain Cognitive Development (ABCD) Study"

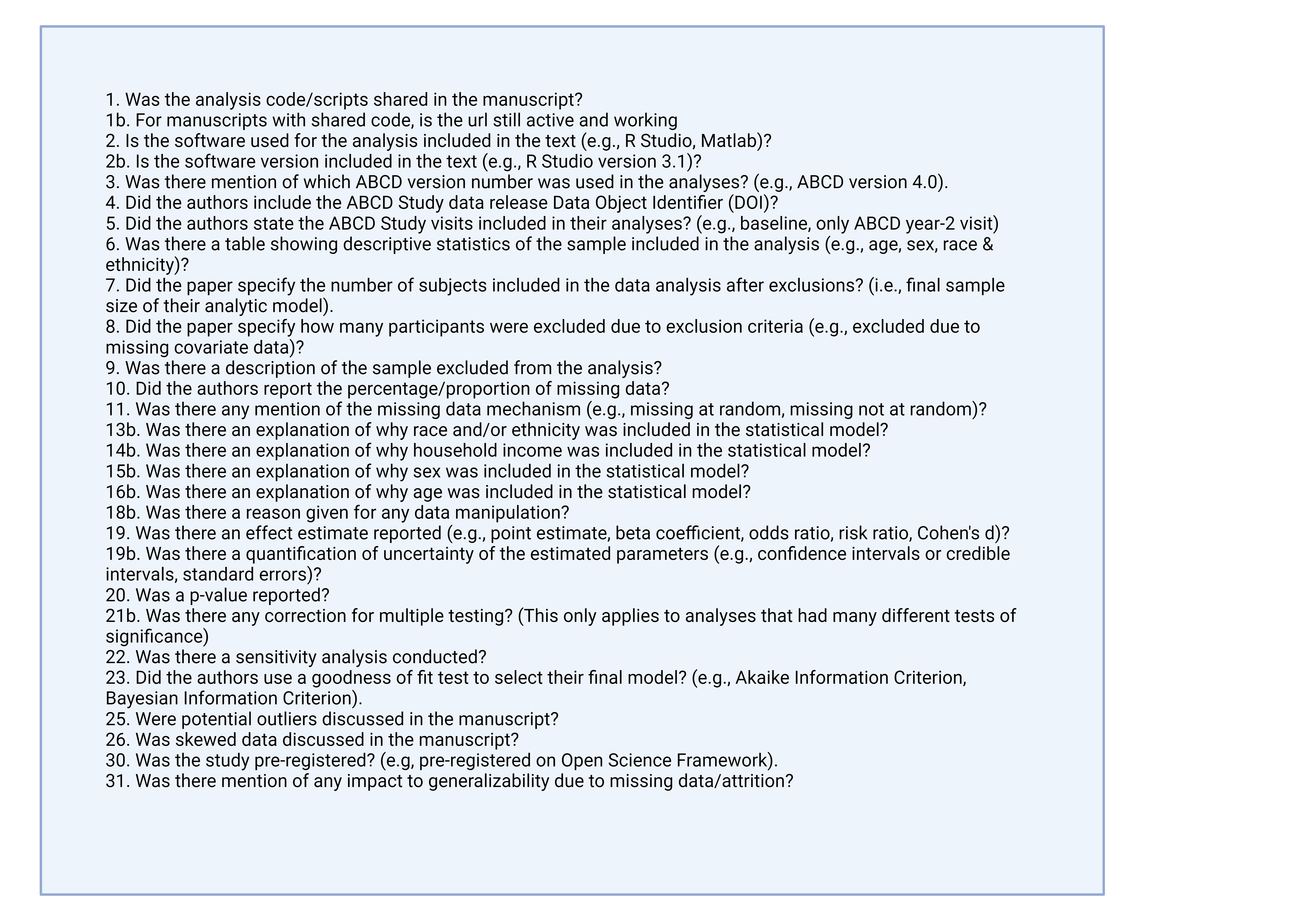

**Figure S1.** Data extraction items comprising the Level of Completeness score

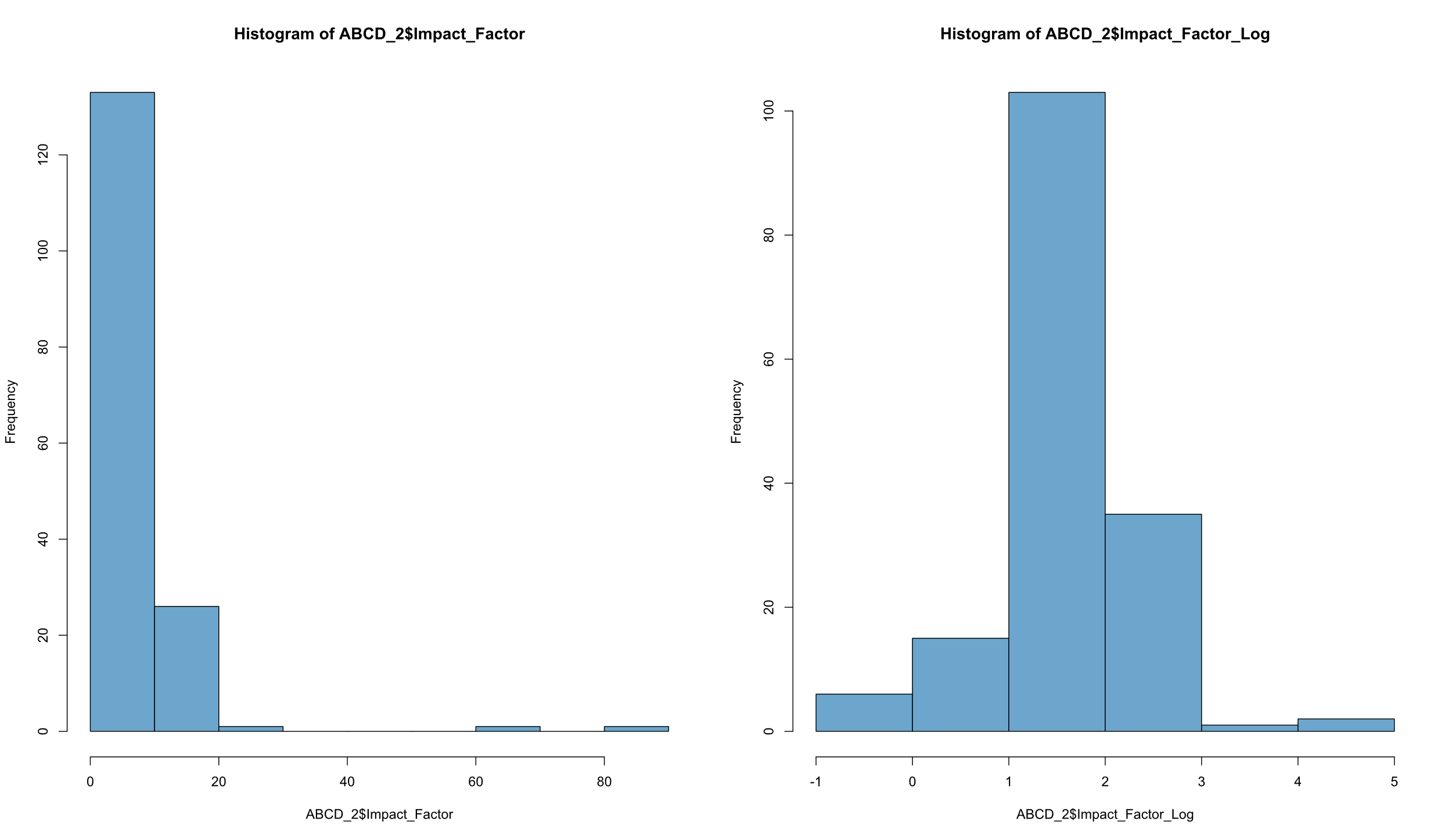

**Figure S2.** (left) histogram of the Impact Factor variable and the (right) logged Impact Factor variable.

**Table S1.** One factor solution for Exploratory Factor Analysis

OBLIMIN ROTATED LOADINGS (* significant at 5% level)

1

________

CODE_SHA	0.144*

CODE_LIN	0.558*

SOFTWARE	0.199*

SOFTWARE	0.280*

ABCD_VER	0.428*

ABCD_DOI	0.211*

ABCD_VIS	0.430*

TABLE_ON	0.296*

NUMBER_I	0.346*

NUMBER_E	0.547*

EXCLUDED	0.266*

PROPORTI	0.556*

MISSING_	0.260*

EXPLANAT	0.708*

EXPLANAT	0.818*

EXPLANAT	0.736*

EXPLANAT	0.720*

REASON_M	0.139

EFFECT_E	0.236*

UNCERTAI	0.019

PVALUE_R	0.148

MULTIPLE	0.009

SENSITIV	0.299*

GOODNESS	0.234*

POTENTIA	0.074

SKEWED_D	0.153*

STUDY_PR	0.195*

GENERALI	0.016

**Table S2.** Two factor analysis

OBLIMIN ROTATED LOADINGS (* significant at 5% level)

1	2

________	________

CODE_SHA	0.223*	-0.033

CODE_LIN	0.461*	0.335

SOFTWARE	0.245*	0.034

SOFTWARE	0.323*	0.090

ABCD_VER	0.598*	0.006

ABCD_DOI	0.275*	0.023

ABCD_VIS	0.515*	0.078

TABLE_ON	0.245*	0.167*

NUMBER_I	0.352*	0.139

NUMBER_E	0.800*	-0.055

EXCLUDED	0.373*	-0.008

PROPORTI	0.775*	-0.013

MISSING_	0.490*	-0.124

EXPLANAT	-0.223*	0.936*

EXPLANAT	0.021	0.858*

EXPLANAT	0.102*	0.840*

EXPLANAT	0.227*	0.758*

REASON_M	0.106	0.102

EFFECT_E	0.285*	0.037

UNCERTAI	-0.174*	0.208*

PVALUE_R	0.218*	-0.020

MULTIPLE	0.166	-0.149

SENSITIV	0.375*	0.067

GOODNESS	0.333*	0.002

POTENTIA	0.258*	-0.158*

SKEWED_D	0.284*	-0.069

STUDY_PR	0.244*	0.014

GENERALI	0.103	-0.095

**Table S3.** Three factor analysis

OBLIMIN ROTATED LOADINGS (* significant at 5% level)

1	2	3

________	________	________

CODE_SHA	0.081	0.012	0.486*

CODE_LIN	0.535*	0.302	-0.190

SOFTWARE	0.271*	0.017	-0.073

SOFTWARE	0.289*	0.093	0.123

ABCD_VER	0.531*	0.013	0.257*

ABCD_DOI	0.277*	0.016	0.015

ABCD_VIS	0.540*	0.058	-0.045

TABLE_ON	0.387*	0.128	-0.399*

NUMBER_I	0.462*	0.099	-0.287*

NUMBER_E	0.840*	-0.090*	-0.031

EXCLUDED	0.372*	-0.016	0.028

PROPORTI	0.733*	-0.020	0.166

MISSING_	0.398*	-0.109	0.309*

EXPLANAT	-0.188*	0.931*	-0.116

EXPLANAT	0.066	0.850*	-0.114

EXPLANAT	0.043	0.852*	0.152*

EXPLANAT	0.150*	0.767*	0.216*

REASON_M	-0.010	0.130	0.316*

EFFECT_E	0.418*	-0.017	-0.379*

UNCERTAI	-0.155*	0.207*	-0.070

PVALUE_R	0.289*	-0.056	-0.214

MULTIPLE	0.225	-0.173	-0.148

SENSITIV	0.287*	0.085	0.291*

GOODNESS	0.233*	0.023	0.330*

POTENTIA	0.194*	-0.147*	0.209*

SKEWED_D	0.207*	-0.054	0.243*

STUDY_PR	0.050	0.076	0.611*

GENERALI	0.094	-0.094	0.037

**Table S4.** Four factor analysis

OBLIMIN ROTATED LOADINGS (* significant at 5% level)

1	2	3	4

________	________	________	________

CODE_SHA	0.184	0.000	0.376	-0.286

CODE_LIN	0.471*	0.315	-0.145	0.189

SOFTWARE	0.200*	0.019	0.013	0.186

SOFTWARE	0.279*	0.092	0.130	0.018

ABCD_VER	0.513*	0.010	0.285*	0.035

ABCD_DOI	0.248*	0.017	0.047	0.077

ABCD_VIS	0.502*	0.065	-0.027	0.129

TABLE_ON	0.381*	0.155	-0.465*	0.139

NUMBER_I	0.335*	0.111	-0.139	0.323*

NUMBER_E	0.822*	-0.077	-0.057	0.099

EXCLUDED	0.524*	-0.005	-0.205	-0.251*

PROPORTI	0.753*	-0.014	0.103	-0.011

MISSING_	0.518*	-0.112	0.132	-0.256*

EXPLANAT	-0.139*	0.940*	-0.185*	-0.124

EXPLANAT	0.109	0.842*	-0.170*	-0.071

EXPLANAT	-0.010	0.849*	0.219*	0.089

EXPLANAT	0.075	0.762*	0.333*	0.139

REASON_M	-0.023	0.119	0.366*	-0.019

EFFECT_E	0.166	-0.012	-0.045	0.659*

UNCERTAI	-0.197*	0.205*	-0.002	0.080

PVALUE_R	0.076	-0.051	0.086	0.470*

MULTIPLE	0.001	-0.172	0.191	0.476*

SENSITIV	0.283*	0.078	0.311*	-0.012

GOODNESS	0.218*	0.012	0.381*	0.004

POTENTIA	0.185*	-0.153*	0.238*	0.005

SKEWED_D	0.147	-0.065	0.360*	0.110

STUDY_PR	0.243	0.067	0.377	-0.486

GENERALI	0.198*	-0.090	-0.128	-0.206*

| **Table S5.** Regression results with Extraction Items and Level of Completeness Score | | |  |
| --- | --- | --- | --- |
|  | **Level of Completeness score** |  |  |
|  | **β (95% CI)** | **t score** | ***p*-value** |
| **Measure** |  |  |  |
| **Year of Publication** | 0.83 (0.55, 1.11) | 5.8 | 9.5×10^-9^ |
| **Imaging Data Used** |  |  |  |
| Yes | 0.6 (-0.06, 1.25) | 1.8 | 0.075 |
| **Genetics Data Used** |  |  |  |
| Yes | 0.52 (-0.38, 1.41) | 1.1 | 0.26 |
| **Machine learning methods** | |  |  |
| Yes | -0.45 (-1.45, 0.55) | -0.88 | 0.38 |
| **Code Shared** |  |  |  |
| Yes | 3.04 (2.37, 3.7) | 8.91 | 2×10^-16^ |
| **Code Link Working** |  |  |  |
| Yes | 3.46 (1.2, 5.7) | 3.02 | 0.003 |
| **Software Used** |  |  |  |
| Yes | 3.38 (2.3, 4.5) | 6 | 3.7×10^-9^ |
| **Software Version** |  |  |  |
| Yes | 2.6 (1.9, 3.3) | 7 | 1.03×10^-11^ |
| **ABCD Version** |  |  |  |
| Yes | 3.8 (3.1, 4.6) | 10.2 | 2×10^-16^ |
| **ABCD DOI** |  |  |  |
| Yes | 2.31 (1.7, 2.94) | 7.2 | 2.1×10^-12^ |
| **ABCD Visits** |  |  |  |
| Yes | 3.4 (2.6, 4.2) | 8.7 | 2×10^-16^ |
| **Demographics Table** |  |  |  |
| Yes | 2.7 (1.9, 3.6) | 6.5 | 2.2×10^-10^ |
| **Data Manipulation** |  |  |  |
| Yes | 2.8 (2.14, 3.37) | 8.8 | 2×10^-16^ |
| **Reason for Manipulation** |  |  |  |
| Yes | 1.5 (0.65, 2.33) | 3.5 | 0.0005 |
| **Effect Estimate Reported** |  |  |  |
| Yes | 3.5 (2.7, 4.4) | 8.5 | 2×10^-16^ |
| **Uncertainty Parameter** |  |  |  |
| Yes | 0.63 (-0.31, 1.6) | 1.32 | 0.19 |
| **Excluded Demographics (Ref=No)** | |  |  |
| No participants excluded | -2.16 (-3.3, -1.1) | -3.9 | 0.0001 |
| Yes | 2.51 (1.9, 3.2) | 7.7 | 8,4×10^-14^ |
| **Explanation for Age** |  |  |  |
| Yes | 3.17 (2.5, 3.8) | 9.7 | 2×10^-16^ |
| **Explanation for Household Income** | |  |  |
| Yes | 2.85 (2.13, 3.6) | 7.8 | 6×10^-14^ |
| **Explanation for Race and/or Ethnicity** | |  |  |
| Yes | 1.62 (0.9, 2.33) | 4.4 | 1.2×10^-5^ |
| **Explanation for Sex** |  |  |  |
| Yes | 2.7 (2.05, 3.4) | 8.1 | 4×10^-15^ |
| **Generalizability Limitations** | |  |  |
| Yes | 2 (1.2, 2.9) | 4.6 | 4.8×10^-6^ |
| No missing data/attrition | -2.5 (-3.5, -1.6) | -5.3 | 2.1×10^-7^ |
| **Goodness of fit test** |  |  |  |
| Yes | 2.6 (1.9, 3.3) | 7.3 | 8.3×10^-13^ |
| **Data Imputation** |  |  |  |
| Yes | 1.9 (1.14, 2.6) | 5 | 9×10^-7^ |
| **Multiple Comparisons Mentioned** | |  |  |
| Yes | 2.4 (1.8, 3.0) | 7.5 | 3.2×10^-13^ |
| **Multiple Comparisons Corrected** | |  |  |
| Yes | 1.4 (-0.05, 2.8) | 1.9 | 0.06 |
| **Missingness Mechanism** |  |  |  |
| Yes | 3.11 (2.3, 4) | 7.2 | 2.1×10^-12^ |
| **N Included in Analytic Sample** | |  |  |
| Yes | 3.5 (2.6, 4.3) | 8.1 | 4×10^-15^ |
| **N Excluded** |  |  |  |
| No participants excluded | -0.61 (-1.6, 0.42) | -1.2 | 0.243 |
| Yes | 4.2 (3.6, 4.8) | 13.3 | 2×10^-16^ |
| **P-value Reported** |  |  |  |
| Yes | 2.9 (1.8, 3.9) | 5.2 | 2.8×10^-7^ |
| **Potential Outliers** |  |  |  |
| Yes | 2.2 (1.4, 3.1) | 5.4 | 1.3×10^-7^ |
| **Skewed Data** |  |  |  |
| Yes | 2.8 (1.8, 3.7) | 5.9 | 6.6×10^-9^ |
| **Proportion Missing Data** |  |  |  |
| Yes | 4.1 (3.5, 4.7) | 13.63 | 2×10^-16^ |
| **Sensitivity Analysis** |  |  |  |
| Yes | 3.3 (2.64, 3.9) | 10.14 | 2×10^-16^ |
| **Study Pre-registered** |  |  |  |
| Yes | 2.7 (1.52, 3.8) | 4.5 | 6.7×10^-6^ |
| **DEAP Used** |  |  |  |
| Yes | -1.9 (-3.1, -0.72) | -3.2 | 0.0017 |
| **Author Contributions** |  |  |  |
| Yes | 1.04 (0.38, 1.7) | 3.11 | 0.002 |
| **Behavioral only (Ref=No)** |  |  |  |
| Yes | -0.14 (-1.3, 1.0) | -0.24 | 0.808 |
| Note: Model results are from a univariate linear regression analysis | | | |

| **Table S6.** Attribute statistics from the Boruta selection method. | | |  |  |  |  |
| --- | --- | --- | --- | --- | --- | --- |
| **Extraction Item** | **Mean Importance** | **Median Importance** | **Minimum Importance** | **Maximum Importance** | **Normalized Hits** | **Final Decision** |
| Author_Contributions | 0.958529924 | 1.051155992 | -0.38945264 | 3.221316221 | 0.001194268 | Rejected |
| Machine_Learning | 0.629900067 | 0.414951111 | -0.550310903 | 2.045362827 | 0 | Rejected |
| Year_Published | 6.524858773 | 6.559180822 | 3.125110352 | 9.281804394 | 1 | Confirmed |
| Age_Included | 3.159512468 | 3.186828883 | -0.2682916 | 6.074242235 | 0.753582803 | Confirmed |
| Sex_Included | 4.770945086 | 4.811861399 | 1.132709367 | 7.181638768 | 0.984474522 | Confirmed |
| HHIncome_Included | 4.309090747 | 4.326159352 | 1.159271801 | 7.779537954 | 0.962181529 | Confirmed |
| Race_Included | 3.670093407 | 3.679362618 | 0.608226304 | 6.589409745 | 0.871417197 | Confirmed |
| Study_PreRegistered | 7.053726451 | 7.059140486 | 4.353874418 | 9.502712352 | 1 | Confirmed |
| Genetics_Used | 2.543918289 | 2.55567772 | -0.715785158 | 5.256235652 | 0.544984076 | Confirmed |
| ROI_Rationale | 2.526531038 | 2.520150424 | -0.510185844 | 6.050724348 | 0.535031847 | Confirmed |
| Imaging_Used | 3.482130797 | 3.495651469 | -0.615729582 | 6.556074167 | 0.822452229 | Confirmed |
| Skewed_Data | 7.9218902 | 7.938602847 | 4.897055169 | 11.10835699 | 1 | Confirmed |
| Potential_Outliers | 8.54800359 | 8.557230043 | 5.768928755 | 11.09029252 | 1 | Confirmed |
| DEAP_Used | 2.53504456 | 2.513233175 | -0.851296926 | 5.979271423 | 0.538216561 | Confirmed |
| Data_Imputation | 6.981586606 | 6.983052705 | 3.966480386 | 9.222975046 | 1 | Confirmed |
| Goodness_Fit | 10.30513543 | 10.30215989 | 7.527889881 | 13.61504012 | 1 | Confirmed |
| Sensitivity_Analysis | 16.48279068 | 16.48076327 | 13.88566588 | 19.88295944 | 1 | Confirmed |
| Multiple_Correction | 10.41658502 | 10.41014983 | 7.220379536 | 12.89674678 | 1 | Confirmed |
| Multiple_Comparisons | 8.483049223 | 8.491945124 | 5.612364615 | 11.16475117 | 1 | Confirmed |
| Pvalue_Reported | 3.87772351 | 3.894452542 | 0.801242497 | 7.052865796 | 0.899681529 | Confirmed |
| Uncertainty_Parameter | 14.41281042 | 14.40554478 | 11.70314482 | 17.03522674 | 1 | Confirmed |
| Effect_EstimateReported | 12.66943825 | 12.67446608 | 10.53822209 | 15.13244292 | 1 | Confirmed |
| Reason_Manipulation | 12.57342548 | 12.5805883 | 9.837250482 | 15.35379368 | 1 | Confirmed |
| Data_Manipulation | 10.38204631 | 10.41314926 | 7.391894678 | 12.57378789 | 1 | Confirmed |
| Explanation_Sex | 16.43760053 | 16.40080407 | 13.75873646 | 19.20830426 | 1 | Confirmed |
| Explanation_Race | 12.6739909 | 12.6568994 | 9.983920783 | 15.2165286 | 1 | Confirmed |
| Explanation_HHIncome | 16.97634372 | 16.97761456 | 14.31725196 | 20.3261202 | 1 | Confirmed |
| Explanation_Age | 17.46198127 | 17.46013063 | 14.96294421 | 20.25232482 | 1 | Confirmed |
| Missing_Mechanism | 10.12477544 | 10.1428054 | 6.488388549 | 12.33155045 | 1 | Confirmed |
| Proportion_MissingData | 20.69332116 | 20.66258915 | 17.70709099 | 23.5409258 | 1 | Confirmed |
| Excluded_Demographics | 13.75122251 | 13.73393287 | 11.18505709 | 16.35765126 | 1 | Confirmed |
| Number_Excluded | 23.86270647 | 23.81566318 | 21.27538951 | 26.70933975 | 1 | Confirmed |
| Number_Included | 9.025136809 | 9.014293067 | 6.403686724 | 11.15149932 | 1 | Confirmed |
| Table_One | 8.765100161 | 8.761903457 | 5.65591751 | 11.69307083 | 1 | Confirmed |
| ABCD_Visits | 11.7153593 | 11.70325234 | 9.565988487 | 13.75144325 | 1 | Confirmed |
| ABCD_DOI | 10.79775555 | 10.78883866 | 8.246653823 | 13.43053736 | 1 | Confirmed |
| ABCD_Version | 15.21399533 | 15.19625901 | 12.87895778 | 17.84599629 | 1 | Confirmed |
| Software_Version | 14.93764395 | 14.94524106 | 12.56889186 | 17.57383296 | 1 | Confirmed |
| Software_Used | 7.846318519 | 7.867002281 | 5.034909447 | 10.07715221 | 1 | Confirmed |
| Code_Shared | 14.93064393 | 14.93558579 | 12.31260208 | 17.36341246 | 1 | Confirmed |
| Code_Link | 16.27552244 | 16.29090774 | 13.23943828 | 18.98865099 | 1 | Confirmed |

| **Table S7.** The correlation between Completeness scores and logged impact factor | | |
| --- | --- | --- |
|  | **Impact Factor (Log)** | |
|  | **β** | ***p*-value** |
| **Level of Completeness (reference=High)** | |  |
| Middle 10% (n=51) | -0.08 (-0.34, 0.18) | 0.537 |
| Bottom 10% (n=52) | -0.77 (-1.02, -0.51) | 2.6×10^8^ |
| **Level of Completeness (continuous) (n=158)** | 0.056 (0.037, 0.074) | 2.3×10^8^ |

**Note.** Estimates are the result of a univariate linear regression

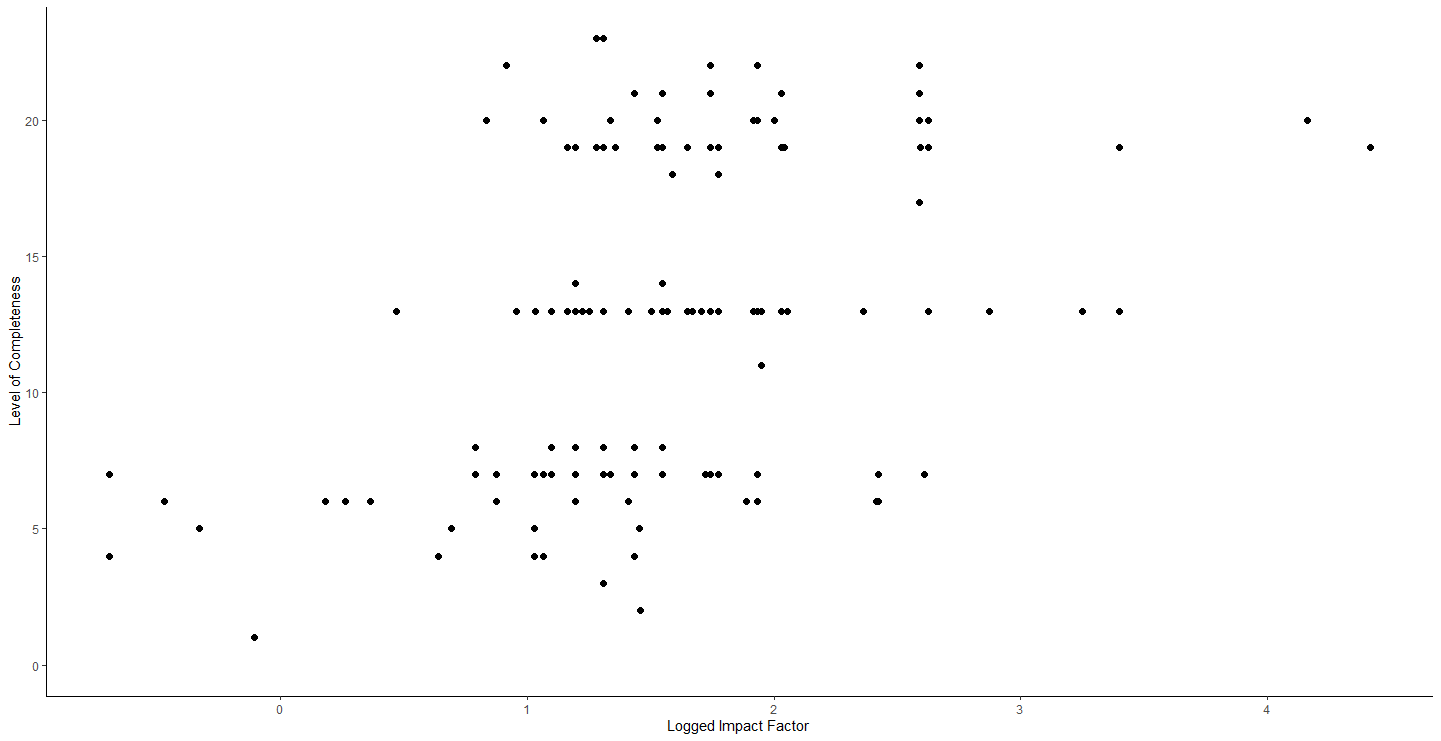

**Figure S3.** A scatter plot of the logged impact factor (x-axis) and the Level of Completeness score (y-axis).

**Supplement Methods**

**The following survey items were developed based on previously existing guidelines in neuroimaging studies reporting, and previously generated guidelines for researchers developed by the ABCD Justice, Equity, Diversity, and Inclusion – Responsible Use of Data Workgroup.**

**Klapwijk, E. T., van den Bos, W., Tamnes, C. K., Raschle, N. M., & Mills, K. L. (2021). Opportunities for increased reproducibility and replicability of developmental neuroimaging. Developmental Cognitive Neuroscience, 47, 100902. https://doi.org/10.1016/j.dcn.2020.100902**

**Nichols, T. E., Das, S., Eickhoff, S. B., Evans, A. C., Glatard, T., Hanke, M., Kriegeskorte, N., Milham, M. P., Poldrack, R. A., Poline, J.-B., Proal, E., Thirion, B., Van Essen, D. C., White, T., & Yeo, B. T. T. (2017). Best practices in data analysis and sharing in neuroimaging using MRI. Nature Neuroscience, 20(3), 299–303. https://doi.org/10.1038/nn.4500**

**Bodison, S. C., Nagel, B., Lopez, D. A., Huber, R., & Members of ABCD JEDI WG3. (2023). Equity-Focused Questions for Researchers using the ABCD Study. https://osf.io/pm7sy/**

**Survey items**

1. Was the analysis code/scripts shared in the manuscript? For example, is there a digital object identifier (DOI) or URL link that takes you directly to the code on Github/ Open Science Framework, or some other site?

1b. For manuscripts with shared code, is the url still active and working (i.e., is it archived in a long-term accessible location on the web)?

2. Is the software used for the analysis included in the text (e.g., R Studio, Matlab)?

2b. Is the software version included in the text (e.g., R Studio version 3.1)?

3. Was there mention of which ABCD version number was used in the analyses? (e.g., ABCD version 4.0).

4. Did the authors include the ABCD Study data release Data Object Identifier (DOI)?

5. Did the authors state the ABCD Study visits included in their analyses? (e.g., baseline, only ABCD year-2 visit)

6. Was there a table showing descriptive statistics of the sample included in the analysis (e.g., age, sex, race & ethnicity)?

7. Did the paper specify the number of subjects included in the data analysis after exclusions? (i.e., final sample size of their analytic model).

8. Did the paper specify how many participants were excluded due to exclusion criteria (e.g., excluded due to missing covariate data)?

9. Was there a description of the sample excluded from the analysis?

10. Did the authors report the percentage/proportion of missing data?

11. Was there any mention of the missing data mechanism (e.g., missing at random, missing not at random)?

12. Did the authors use data imputation methods to account for missing data (e.g., multiple imputation, full information maximum likelihood)?

13. Was there inclusion of race and/or ethnicity in the statistical model?

13b. Was there an explanation of why race and/or ethnicity was included in the statistical model?

14. Was there inclusion of household income in the statistical model?

14b. Was there an explanation of why household income was included in the statistical model?

15. Was there inclusion of participant sex in the statistical model?

15b. Was there an explanation of why sex was included in the statistical model?

16. Was there inclusion of participant age in the statistical model?

16b. Was there an explanation of why age was included in the statistical model?

17. Was there an explanation of why other variables (e.g., parental monitoring) were included in the statistical model?

18. Was there a description of any data manipulation (e.g., changing a variable from continuous to categorical)?

18b. Was there a reason given for any data manipulation?

19. Was there an effect estimate reported (e.g., point estimate, beta coefficient, odds ratio, risk ratio, Cohen's d)?

19b. Was there a quantification of uncertainty of the estimated parameters (e.g., confidence intervals or credible intervals, standard errors)?

20. Was a p-value reported?

21. Did the authors mention issues with multiple comparisons?

21b. Was there any correction for multiple testing? (This only applies to analyses that had many different tests of significance)

22. Was there a sensitivity analysis conducted?

23. Did the authors use a goodness of fit test to select their final model? (e.g., Akaike Information Criterion, Bayesian Information Criterion).

24. Did the paper use the Data Exploration and Analysis Portal (DEAP) to run their analysis?

25. Were potential outliers discussed in the manuscript?

26. Was skewed data discussed in the manuscript?

27. Did the study use machine-learning methods?

28. Was imaging data used in the analysis?

28b. Was there a rationale and method for selecting the particular regions of interest (e.g., did they specify why they included the nucleus accumbens in their analysis)?

29. Was genetics data used in the analysis?

30. Was the study pre-registered? (e.g, pre-registered on Open Science Framework).

31. Was there mention of any impact to generalizability due to missing data/attrition?

32. Did the authors indicate author contributions?
